# Variability in physical inactivity responses of university students during COVID-19 pandemic: A monitoring of daily step counts using a smartphone application

**DOI:** 10.1101/2021.12.21.21268155

**Authors:** Shoji Konda, Issei Ogasawara, Kazuki Fujita, Chisa Aoyama, Teruki Yokoyama, Takuya Magome, Chen Yulong, Ken Hashizume, Tomoyuki Matsuo, Ken Nakata

## Abstract

This study investigated the changes in physical inactivity of university students during the COVID-19 pandemic, with reference to their academic calendar. We used the daily step counts recorded by a smartphone application (iPhone Health App) from April 2020 to January 2021 (287 days) for 603 students. The data for 287 days were divided into five periods based on their academic calendar. The median value of daily step counts across each period was calculated. A k-means clustering analysis was performed to classify the 603 participants into subgroups to demonstrate the variability in the physical inactivity responses. The median daily step counts, with a 7-days moving average, dramatically decreased from 5,000 to 2,000 steps/day in early April. It remained at a lower level (less than 2,000 steps/day) during the first semester, then increased to more than 5,000 steps/day at the start of summer vacation. The clustering analysis demonstrated the variability in physical inactivity responses. Independent of the academic calendar, many inactive students did not recover their original daily step counts after its dramatic decrement. Consequently, promoting physical activity is recommended for inactive university students over the course of the whole semester.

## INTRODUCTION

The Coronavirus disease (COVID-19) has severely affected daily life activities around the world. Governments worldwide have enacted restrictive measures to reduce the risk of infection. The restrictions have led to physical inactivity that increases the risk for non-communicable diseases [1]. Even before the COVID-19 pandemic, physical inactivity had been recognized as a global contributor to the development of chronic non-communicable diseases [2]. The worldwide prevalence of physical inactivity, which does not meet the guideline of World Health Organization (WHO) [3], has been reported to be 27.5% across 168 countries between 2002 and 2016[4]. The COVID-19 pandemic is expected to further accelerate this trend. The influence of the COVID-19 pandemic on physical activity has been reported and summarized in systematic reviews [1,5]. Accordingly, physical activity has been reported to be reduced across all reviewed populations due to the restrictive measures introduced during the COVID-19 pandemic [1]. Furthermore, physical activity has been reported to be associated with lower levels of depression and anxiety during the COVID-19 pandemic [5] and the reduced risk for severe COVID-19 [6-8]. Therefore, it is suggested that physical activity should be evaluated and prescribed, if appropriate, during and after the COVID-19 pandemic.

The daily step counts (sum of steps taken per day) have been considered to be an intuitive metric related to health outcomes monitoring with the high degree of temporal resolution [9,10]. Saint-Maurice et al. demonstrated the importance of daily step counts. Accordingly, adults over 40 years old who recorded more than 8,000 steps/day showed a lower mortality than those who recorded less than 4,000 steps/day [11]. The effectiveness of daily step counts recorded through smartphone applications has been recognized [12].

This metrics has been used to indicate levels of physical activity during the monitoring of large-scale global trends [13]. The survey with the largest scale of daily step counts used a dataset of 717,527 people across 111 countries, recorded through a smartphone application [13]. Smartphone applications have the advantage of high penetration rate, avoiding the need to distribute activity trackers for research. After the COVID-19 pandemic, the sharp decline in the daily step counts in each country coincided with the initiation of restrictive measures by their respective governments [14-20]. The daily step counts gradually recovered during the restrictive measures, after the relaxation of the restrictions, and then increasingly after their removal of the restrictive measures [14-16,19,20]. A survey of daily step counts using a smartphone application can provide real-time information about activity levels on a national or global basis.

During the COVID-19 pandemic, university classes moved online, and university students could not attend classes or extracurricular activities on campus. These developments led to a worldwide decrease in physical activity among university students as is clearly demonstrated by several surveys [21-26]. A study surveyed university students and employees and revealed that light physical activity decreased in undergraduate students but not in others (graduate student, faculty, staff, and administration) [25]. It speculated that these results can be explained by the fact that before the pandemic, undergraduate students walked across campus to attend classes and met faculty and administrators at their offices [25]. The decrease in moderate as well as vigorous physical activity during the pandemic was greater in younger (18–29 years) and older (≥ 79 yrs.) individuals compared with those who are middle-aged [27]. In addition, the daily step counts of participants who normally worked during lockdown in Italy decreased from 11,045 to 5,043 (62% decrease), and for those who remained at home, it decreased from 7,700 to 2,924 (55% decrease) [28]. The physical activity of university students, therefore, was substantially affected by social restrictions associated with the COVID-19 pandemic that has been reported to be a cause of increased depression of university students [26]. It is unclear, however, whether the decreases in physical activity during this period—as observed in these studies—generalize accurately to all students. Changes in physical activity due to the COVID-19 pandemic have been found to be heterogeneous [29,30]. The smartphone-based survey was expected to enable efficient monitoring of the variation of physical activity because most university students already owned smartphones. Therefore, we aimed to investigate the changes in physical inactivity of university students, occurring in response to the COVID-19 pandemic, and as influenced by the academic calendar.

## MATERIALS AND METHODS

### Data collection

The study protocol was approved by the observation research ethics review committee of the Osaka University Hospital (code: 19537). We followed the ethical recommendations for human research as stipulated by the Declaration of Helsinki. We used a daily step counts dataset recorded from students who took Health and Sport Science classes at a national university in Osaka, Japan. An opt-out opportunity was guaranteed for students after the end of the semester. The participants were instructed to use own smartphone devices. According to our preliminary survey, more than 99% of the university students owned smartphones. The participants who used the iPhone and Android devices were recommended to use Apple Health application (Apple Inc.) and Google Fit (Google LCC.), respectively, for educational purpose. There were 913 valid analyzable records and 25 invalid records across participants. Of the 913 valid records, 603 records from the Apple Health application installed on iPhones were used in the study (395 males and 208 females). Google Fit starts to record after installation while Apple Health application starts to record automatically. Thus, the Google Fit users could not record the daily step counts before the start of the first semester. Therefore, we adopted only Apple Health data for research purpose. The measurement period was 287 days, from April 1, 2020 to January 12, 2021.

### Data analysis

The daily step counts were defined as the sum of the steps taken per day. The descriptive statistics were represented by the median, 25% and 75% quantiles for each day across all participants. The moving averages with 7-days (1 week) windows were used to detect the seasonal changes in daily step counts during the investigation period. Colormaps were used to observe inter-subject variation in the seasonal changes in daily step counts. In these colormaps, rows represent individual data ordered from top to bottom by average daily step counts taken over the entire recording period. The colormaps were then filtered using a two-dimensional moving average, with a kernel (10 individuals × 7 days) to describe seasonal and group trends.

The entire recording period was divided into five periods according to the university’s academic calendar as follows: (1) the scheduled preparation period (8 days), defined as the period of guidance that was set to occur immediately after the university’s entrance ceremony, regardless of the COVID-19 pandemic; (2) the unscheduled preparation period (11 days) accommodating the implementation of online classes owing to the COVID-19 pandemic; (3) the first semester (103 days) that ran from spring to summer; (4) the summer vacation (61 days) that followed; (5) and the second semester (104 days) that ran from fall to winter (Figure 1). The cross-correlation matrix with Pearson’s coefficient was used to detect relationships between median daily step counts during the five periods for examining the influence of the academic calendar. Principal component analysis was performed for feature extraction of the median daily step counts during the same five periods. A k-means clustering was performed using principal components and then evaluated using the silhouette value. The number of clusters was determined by the elbow method. The daily step counts withs a 7-days moving averages, were classified based on the results of the k-means clustering.

**Figure 1.**
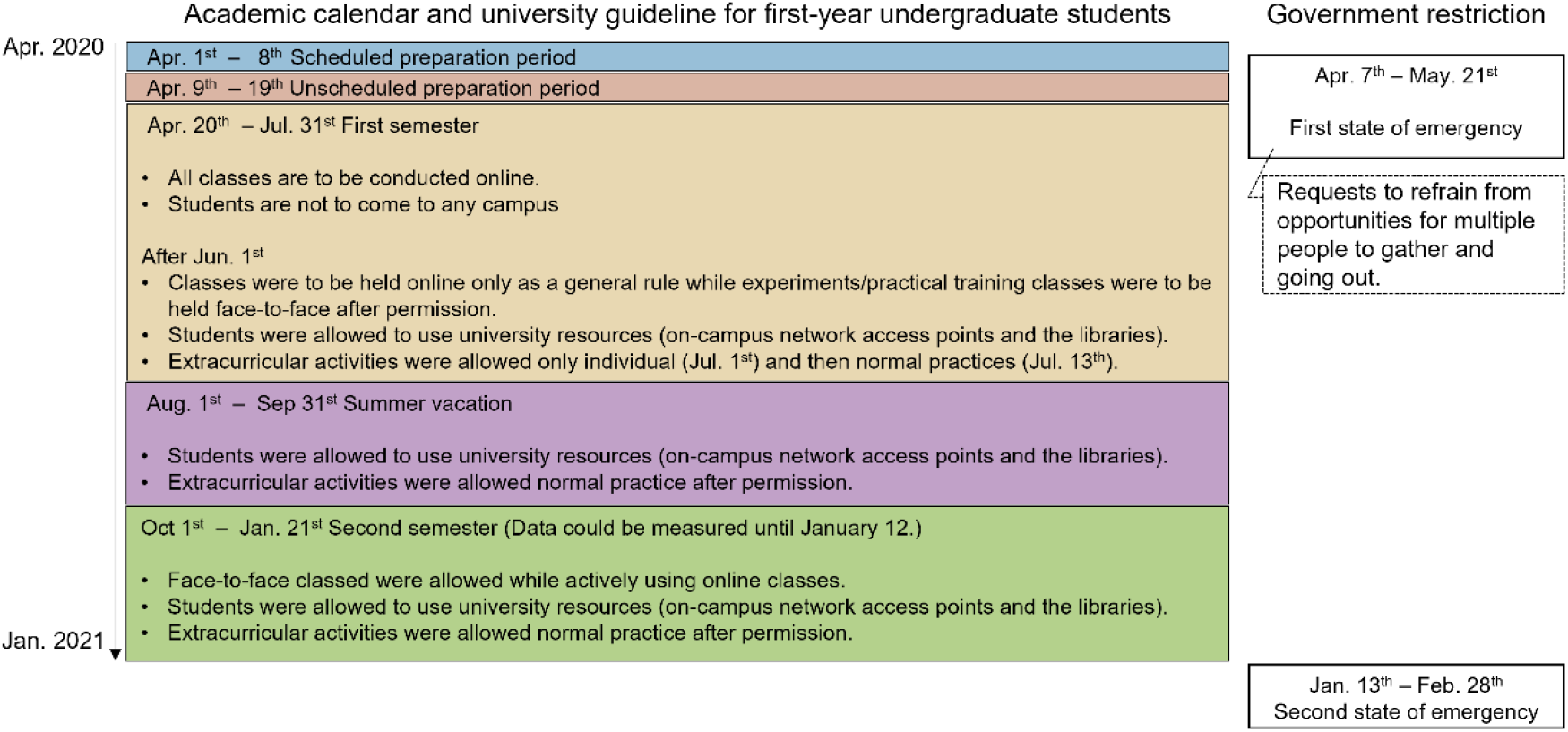
Summary of academic calendar with university guidelines for first-year undergraduate students and Japanese government restrictions.

## RESULTS

Figure 2 shows the time-series changes in median daily step counts with 25% and 75% quantiles across 603 participants. Spikes in daily step counts were recorded at the beginning of April (9,301 steps/day) and in mid-July (7,903 steps/day), coinciding with the days when most participants had opportunities to visit the university (Figure 2-a). The 7-days moving average daily step counts revealed a dramatic decrease during the first 10 days of April (Figure 2-b). The daily step counts then maintained lower values during the state of emergency in Japan (from April 7 to May 5), gradually increasing after the declaration of a state of emergency was lifted (Figure 2-b).

**Figure 2.**
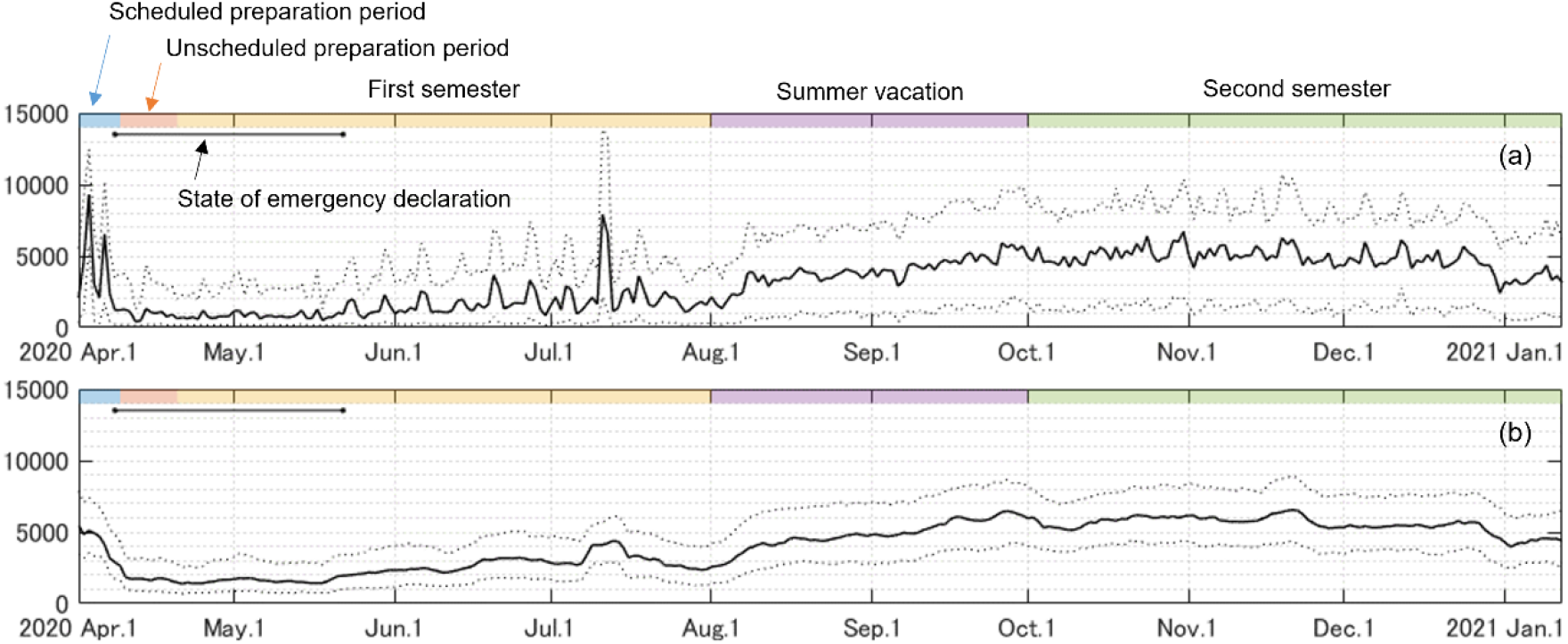
Time-series changes in the median daily step counts. (a) Median daily step counts (solid line) with 25% and 75% quantiles (dotted line) across 603 participants; (b) The 7-days moving average daily step counts (solid line) with 25% and 75% quantiles (dotted line).

In the colormap of the daily step counts (Figure 3-a), each color dot represents the daily step counts of each participant. Participants who recorded over 8,000 mean daily step counts across the target duration are shown with dark red dots, except for the state of emergency. With a decrease in the mean daily step counts (from top to bottom), the blue and dark blue dots increase across the target duration. The colormap of the daily step counts with moving average revealed that the timing of the transition from blue (< 3,000 steps/day) to yellow (> 6,000 steps/day) was delayed backward as the mean daily step counts decreased from top to bottom (Figures 3-b). The transition was small (from blue to light blue) in participants with a lower mean daily step counts (< 2,000) (Figures 3-b).

**Figure 3.**
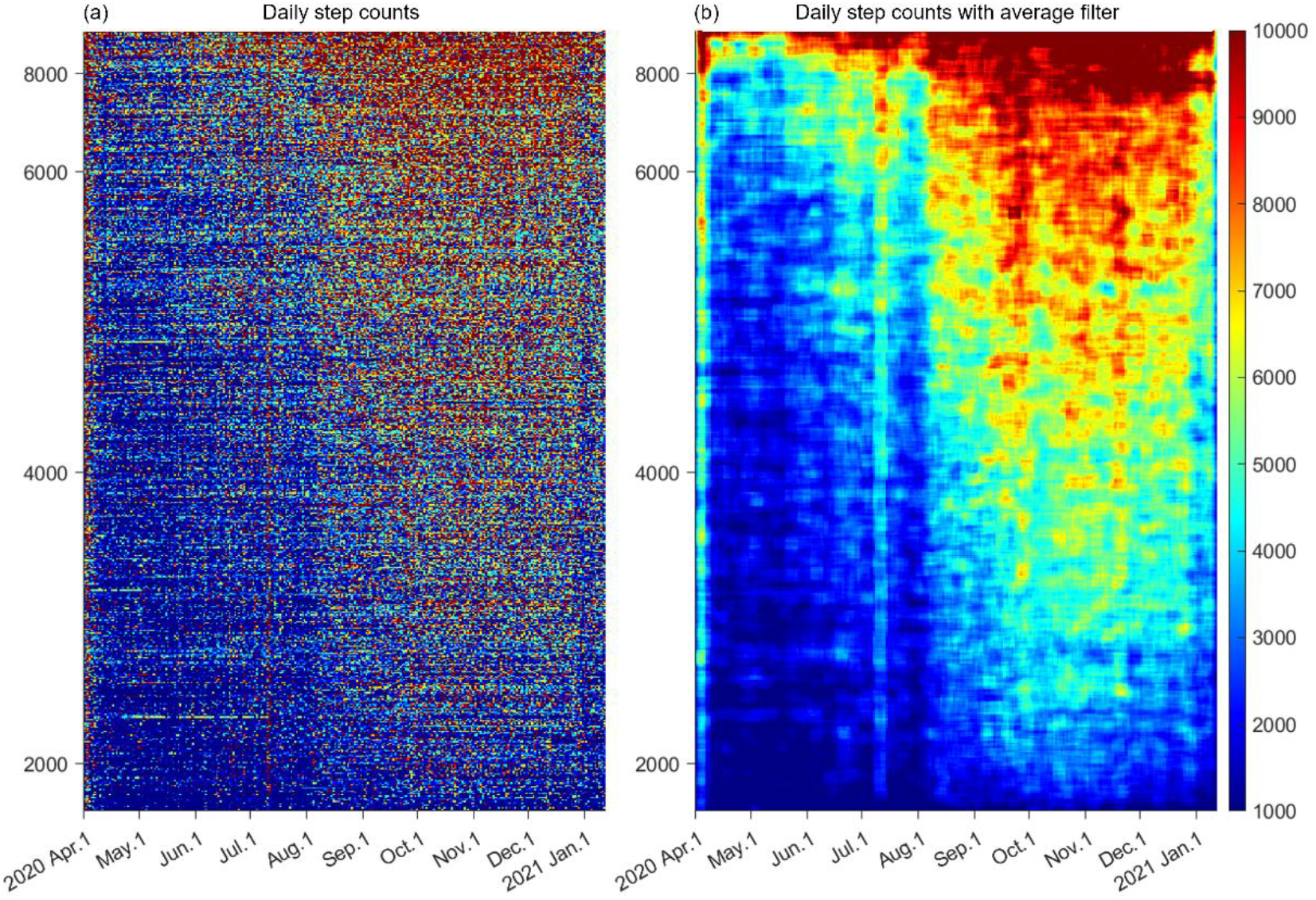
Daily step counts of 603 participants with moving averages. (a)Daily step counts represented as colormaps in descending-order of median daily step counts from top to bottom. (b) Daily step counts after applying average filters with 10 participants × 7 days matrix.

The probabilistic distribution of the mean daily step counts during the five periods— divided according to the academic calendar—were fitted as a gamma distribution (Figure 4). The distribution was extremely skewed toward the lower side (left-skewed) when the median value was relatively low. The lowest median and mode were recorded during the unscheduled preparation period, with a strongly left-skewed distribution. The cross-correlation matrix revealed significant positive correlations for all combinations (p < 0.0001). Relatively strong correlations were observed among the first semester, summer vacation, and the second semester. In contrast, relatively weak correlations were observed for combinations related to scheduled and unscheduled preparation periods. Eighty percent variance was explained by the summation of the first two principal components (Figure 5-a); therefore, the first two principal components were used in the k-means clustering. The number of clusters was set at three, and the silhouette values are presented in Figure 5-b. The distribution of the classified participants on the plane was determined by the two principal components with the vector representing the original variables that shows the daily step counts during five periods according to the academic calendar (Figure 5-c). Figures 5-d, -e, and -f show the time series changes in daily step counts with a 7-days moving average (gray) and the median value across participants in each of the three clusters (red, green, and blue). Clusters 1 (189 participants) and 3 (281 participants) show relatively high and low daily step counts, respectively. Cluster 2 (133 participants) shows a relatively low daily step counts from April to May. The daily step counts then increases starting from June, as shown in clusters 1 and 2. Figure 5 (g) shows the biplot of scores of 603 participants divided in three clusters with loading vectors of five variables on the plane determined by two principal components.

**Figure 4.**
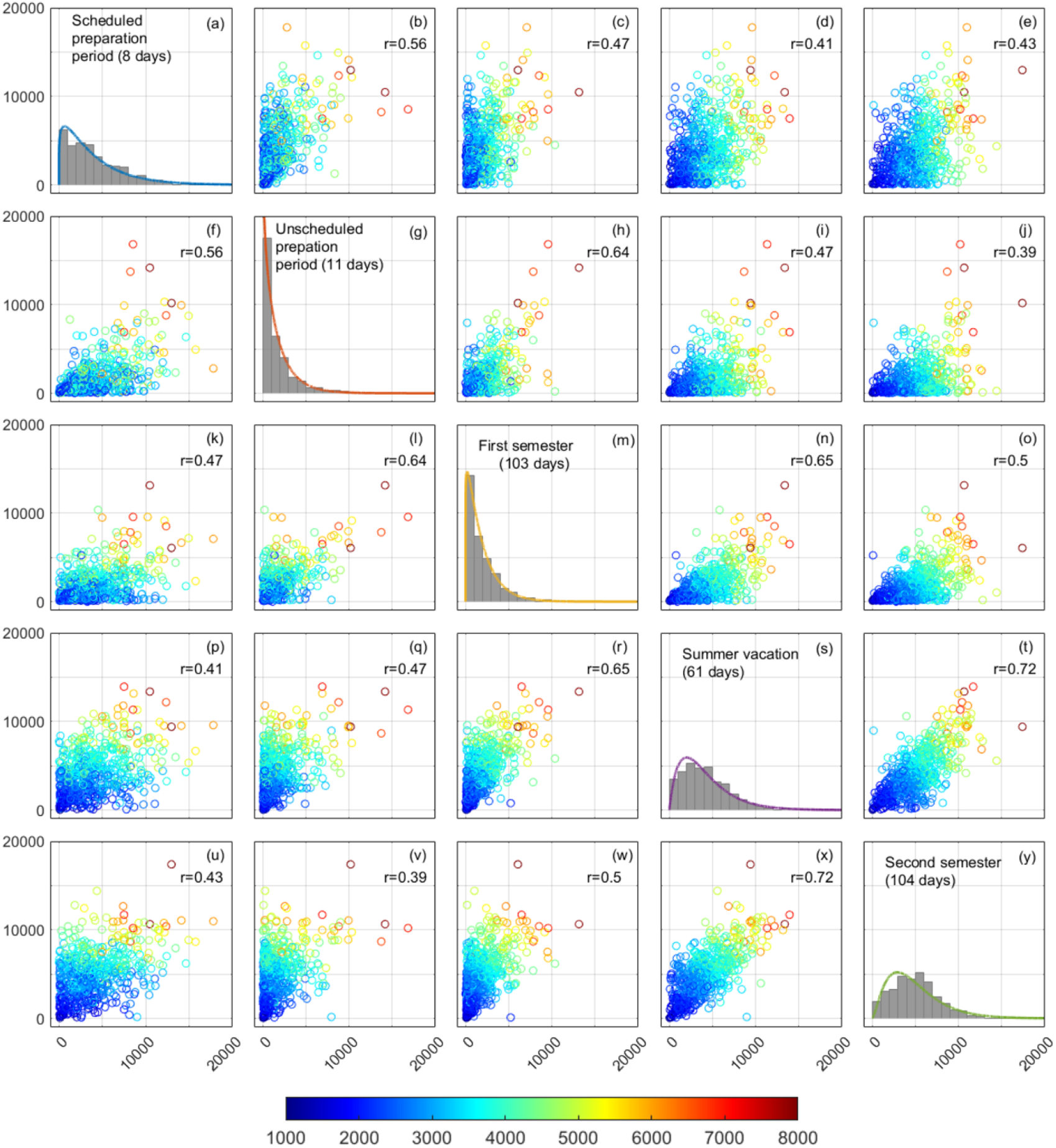
Cross correlation matrix of median daily step counts during five periods according to the academic calendar. Color of scatter represents the median step count of 603 participants during the entire recording period.

**Figure 5.**
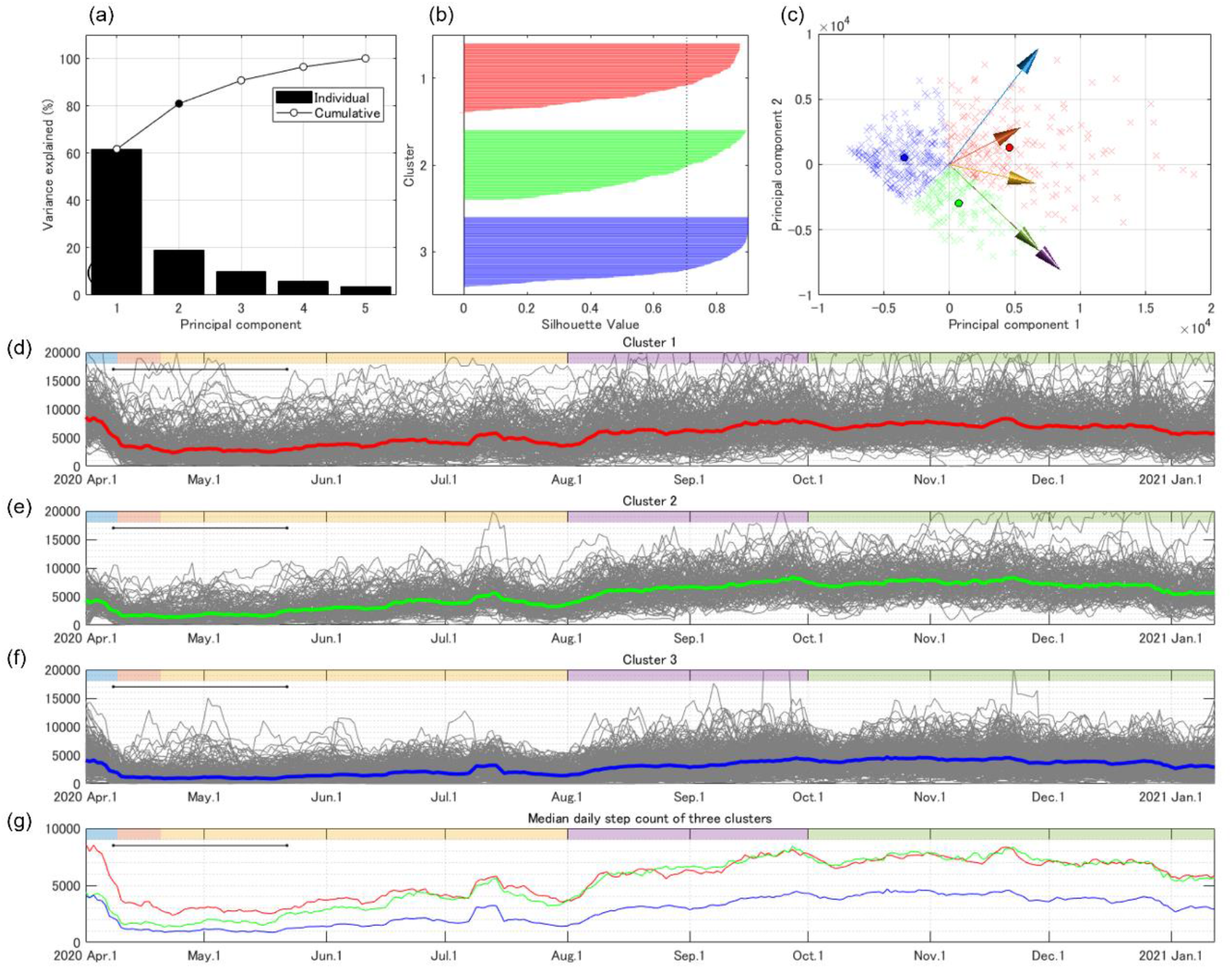
(a) Variance explained by the principal components extracted through the Principal Component Analysis. (b) The accuracy of clustering was evaluated using the silhouette plot. The dotted line shows the mean silhouette value (0.704). (c) Biplot of scores of 603 participants (×) divided into three clusters (red, green, and blue) with the vectors of five original variables (arrows) on the plane determined by two principal components. The colors of the arrows correspond to the colors of each period in Figure 4. Circles showed the mean value for each of the three clusters. (d, e, and f) Time-series changes in the daily step counts with 7-days moving average in three clusters are represented with gray lines, and median values across participants are represented with red (cluster 1), green (cluster 2), and blue (cluster 3). (g) There is a unique pattern of time-series changes between 3 clusters.

## DISCUSSION

We investigated changes in university students’ physical inactivity during the COVID-19 pandemic—according to their academic calendar—by recording daily step counts through smartphone applications. To the best of our knowledge, this is the first study to report the daily step counts of university students across an extended period (287 days), including semester and vacation periods. The high degree of temporal resolution in this study, facilitated by smartphone application use, enabled the detection of continuous changes in the students’ daily step counts. The descriptive results revealed that the daily step counts were also influenced by periods on the academic calendar, as well as socially restrictive governmental measures in response to the COVID-19 pandemic. Additionally, clustering analysis demonstrated variations in the level of physical inactivity owing to the pandemic; for example, many inactive students (47%) did not fully recover from the dramatic decrease in their daily step counts, which remained unaffected by the academic calendar. Physical activity should be promoted for inactive university students— disregarding the academic calendar—during the COVID-19 pandemic.

The daily step counts decreased dramatically at the start of the state of emergency and unscheduled preparation periods in the beginning of April (Figure 2). In global survey data, the mean daily step counts in Japan showed a slow decrease from February to May; and the minimum value (approximately 5,000 steps/day) was observed in mid-April [15]. The baseline mean daily step counts across Japanese participants (N = 20,386) was reported to be 6,010 steps/day in a large-scale global survey using smartphone applications, conducted between July 2013 and December 2014[13]. In data from the nearly age-matched university students before the pandemic of COVID-19 recorded by the activity trackers, the daily step counts were reported to be approximately 7,000 steps/day [31]. From these reports, the rate of decrease owing to the emergency declaration is estimated to be approximately 20%. In contrast, our data showed a reduction of more than 60% before and during the state of emergency. We also compared our daily step counts result (7-days moving average) with the Apple Mobility Trends Reports [32] which well reflects the number of walking bouts as an indicator of physical activity [33]. In this comparison report, we used ‘walking’ mobility trend in Japan. The data were filtered by a 7-days moving average. Also, both the daily step counts and walking mobility trend were normalized using data from April 1, 2020 to compare the rates of decrease. The reduction rate shown in Apple Mobility Trend Reports during the state of emergency was approximately 20–30%, which corresponds to the decrease in daily step counts estimated from previous reports [13,15]. The daily step counts of university students in the current study demonstrated a greater reduction rate and slower recovery than the walking mobility trend in Japan (Figure 6). Likewise, the results of a global survey reported that the daily step counts of a younger group was lower than that of the older group [19,27], as member of younger group remained at home for longer periods during COVID-19 [19]. Thus, the physical activity of university students may be more susceptible to restrictive measures than indicated by the overall trend in Japan, as is indicated in our study. The rate of decline in physical activity of university students in the current study is estimated to be similar to that observed in countries where the government declared strict restrictive measures [14,19,27].

**Figure 6.**
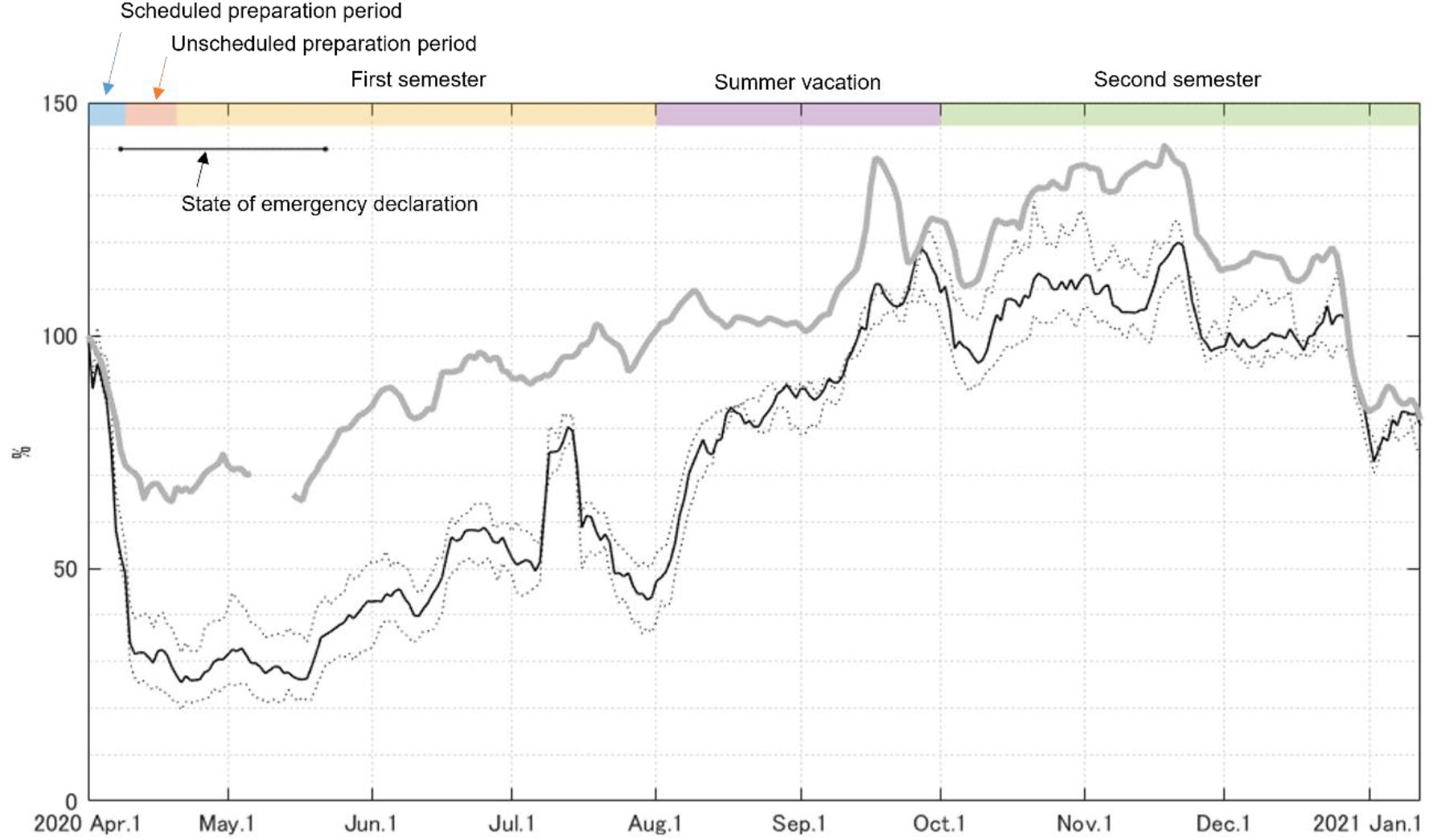
Apple Mobility Trend Reports (Gray solid line) and median daily step counts with 7-days moving average (black solid line) normalized by these data obtained on April 1, 2020.

The probabilistic distribution of the median daily step counts across each period showed unique features (Figures 4-a, -g, -m, -s, and -y). The unscheduled preparation period was extended into the scheduled period to lengthen the time allotted to shifting into online instructional mode. The unscheduled period started at almost the same time as the state of emergency in Japan. The distribution of the median daily step counts is left-skewed, indicating that most participants stayed at home, while a few participants maintained physical activity levels of more than 8,000 steps/day. The left-skewed distribution was slightly corrected during the first semester, but the daily step counts was low (Figure 4-m). Thus, it seems that in this phase, participants who restarted physical activity gradually increased with great individual variations in start date and daily step counts (Figures 3-b). We speculate that most participants did not take time for physical activities between managing online classes and homework during the first semester, even after the end of the state of emergency. Other participants, however, began to make time for physical activity. After the end of the first semester, the daily step counts increased during summer vacation (Figures 2 and 3), and the left-skewed distribution was corrected by a reduction of low daily step counts (Figure 4-s). In the second semester, the university adopted a hybrid of face-to-face and online classes. The median daily step counts reached 6,000 steps/day (Figure 4-y), which is the reported baseline of Japanese daily step counts on average, despite not all participants having opportunities to go to the university campus during the second semester [13]. The contribution of conducting face-to-face classes on physical activity can be observed on days when most students were most likely to go to the university, at the beginning of April and middle of July. The median values across participants exceeded 6,000 steps/day, the 75% quantile exceeded 10,000 steps/day (Figures 1-a) and the 7-days moving averages approached 6,000 and 8,000 steps/day, respectively (Figures 1-b). Therefore, university students are able to meet the recommended daily step counts (8,000 steps/day) when they have opportunities to attend university in-person. It is speculated that over 8,000 steps/day is achieved by commuting to school and walking around the campus. This observation matches previous reports in which light physical activity decreased only in undergraduate students but not in the others who attend or work at the university [25]. The levels of physical activity in university students are thus strongly related to the academic calendar and the decrease in restrictive governmental measures.

There was great variability in the daily step counts among the participants (Figures 3 and 4). The participants with over 8000 steps/day on the median daily step counts of all target periods demonstrated a nearly constant high daily step counts throughout the year (Figures 3 and 4). We speculate that these participants did not dramatically decrease their voluntary physical activity during the unscheduled preparation period and first semester, despite the lack of opportunities to go to university. In contrast, the participants who achieved less than 2,000 steps/day on average during all target periods showed a dramatic decrease in daily step counts during the unscheduled preparation period and first semester, followed by lower degrees of recovery during summer vacation and second semester (Figures 3 and 4). These participants are probably easily influenced by the opportunities to go to university because they do not regularly exercise on their own. The unique features observed in this descriptive visualization (Figures 3, and 4) were quantitatively verified by the k-means clustering (Figure 5). The daily step counts of cluster 2 (green, 22% of participants) (Figure 5-e) shows the lower level of daily step counts from April to mid-May 2020 and the daily step counts increased to the same level as that of cluster 1 (red, 31% of participants) (Figure 5-d). The biplot showed that boundary of cluster 1 and 2 was orthogonal to the vector representing the daily step counts during the scheduled and unscheduled preparation periods (blue and orange) (Figure 5-c). It demonstrated that the major difference between two clusters is the daily step counts during the scheduled and unscheduled preparation periods. The mean daily step counts taken by participants in cluster 2 began to increase, which coincided with the lifting of the state of emergency declaration in mid-May. Cluster 3 (47% of participants) represents the lower level of daily step counts throughout the first semester (Figure 5-f). The biplot showed that boundary of cluster 2 and 3 was orthogonal to the vector representing the daily step counts during the summer vacation (purple) and second semester (green) (Figure 5-c). It demonstrated that the major difference between two clusters is the daily step counts during the summer vacation and second semester. The increase at the start of summer vacation of cluster 3 was smaller than that of clusters 1 and 2. The difference in the daily step counts after August was greater than that earlier in the year. There were many inactive students who remained at a lower level of daily step counts, unaffected by the academic calendar. The low level of daily step counts in cluster 3 (47% of participants) could be due to the group being originally less active, or it could be due to the COVID-19 pandemic, or both. Promoting physical activity is required for inactive university students, regardless of the academic calendar, during the COVID-19 pandemic.

A disadvantage of using a smartphone application is the accuracy of recording step counts in free-living conditions. Smartphone applications enable the recording of continuous steps in the experimental condition where the target step count is set [34,35]. However, the accuracy of step counts, however, recorded in free-living conditions has been known to be underestimated by non-recorded steps that occur when the participant did not hold the smartphone [36]. The major source of recorded daily step counts by smartphones is locomotive activity. In contrast, the step counts cannot be recorded during household and sports activities, resulting in the underestimation of daily step counts in the free-living condition. Most sports activities on and off campus were restricted from April to July 2020, and then permitted from August 2020 onward. The recorded daily step counts accurately represents the physical activity during the scheduled preparation period, unscheduled preparation period, and first semester, but it may underestimate physical activity during summer vacation and second semester. Therefore, we speculated that the influence of the academic calendar on daily step counts, as a major finding of this study, is not impacted by the reported underestimation of the daily step counts recorded by smartphone applications [34-37].

## Conclusion

Our study aimed to examine the variations of physical inactivity during the COVID-19 pandemic, with respect to the academic calendar of the participating students. The smartphone-based survey allowed efficient monitoring of physical activity with high temporal resolution, because more than 99% of university students own smartphones. Our data shows that the daily step counts is greatly influenced by the university academic calendar and governmental restrictive measures in response to the COVID-19 pandemic. Clustering analysis demonstrated that variations in level of physical inactivity occurred, and there were many inactive students (47%) who did not recover their daily step counts after its dramatic decrease, which remained unaffected by the academic calendar. We suggest that the promotion of physical activity is essential to preserve the health of inactive university students as the COVID-19 pandemic continues or similar crises occur, when students take online classes and have no opportunities to travel to the university campus. Long term continuous monitoring of daily step counts in university students is needed to investigate the physical activity during/after the COVID-19 pandemic and impact of increasing online class.

## Data Availability

All data produced in the present study are available upon reasonable request to the authors

## Acknowledgements

We appreciate the generous cooperation of secretaries of Department of Health and Sport Sciences, Osaka University for providing assistance with data collection.

## Author contribution

Conceptualization, S.K., I.O., K.F., and K.N.; Data acquisition, S.K., I.O., K.F., C.A., T.Y., T.M., C.Y., K.H., T.M., and K.N.; Data analysis, S.K., and I.O.; Interpretation, S.K., I.O., K.F., C.A., T.Y., T.M., C.Y., K.H., T.M., and K.N. Writing original draft, S.K. I.O., and K.N. All authors critically revised the report, commented on drafts of the manuscript, and approved the final report.

## Institutional Review Board Statement

The study was conducted in accordance with the guidelines of the Declaration of Helsinki and approved by the Institutional Ethics Committee of Osaka University Hospital (19537-2, July 1, 2020).

## Informed Consent Statement

We used a daily step counts dataset recorded from university students who took Health and Sport Science classes. An opt-out opportunity was guaranteed for the students after the end of the semester for the use of dataset to research purpose.

## Data Availability Statement

The data analyzed in this manuscript will be made available from the corresponding author upon reasonable request.

